# Model-estimated relationship between elementary school-related SARS-CoV-2 transmission, mitigation interventions, and vaccination coverage across community incidence levels

**DOI:** 10.1101/2021.08.04.21261576

**Authors:** John Giardina, Alyssa Bilinski, Meagan C. Fitzpatrick, Emily A. Kendall, Benjamin P. Linas, Joshua Salomon, Andrea L. Ciaranello

## Abstract

**Background:** While CDC guidance for K-12 schools recommends indoor masking regardless of vaccination status, final decisions about masking in schools will be made at the local and state level. The impact of the removal of mask restrictions, however, on COVID-19 outcomes for elementary students, educators/staff, and their households is not well known.

**Methods:** We used a previously published agent-based dynamic transmission model of SARS-CoV-2 in K-12 schools to simulate an elementary school with 638 students across 12 scenarios: combinations of three viral infectiousness levels (reflecting wild-type virus, alpha variant, and delta variant) and four student vaccination levels (0%, 25%, 50% and 70% coverage). For each scenario, we varied observed community COVID-19 incidence (0 to 50 cases/100,000 people/day) and mitigation effectiveness (0-100% reduction to in-school secondary attack rate), and evaluated two outcomes over a 30 day period: (1) the probability of at least one in-school transmission, and (2) average increase in total infections among students, educators/staff, and their household members associated with moving from more to less intensive mitigation measures.

**Results:** Over 30 days in the simulated elementary school, the probability of at least one in-school SARS-CoV-2 transmission and the number of estimated additional infections in the immediate school community associated with changes in mitigation measures varied widely. In one scenario with the delta variant and no student vaccination, assuming that baseline mitigation measures of simple ventilation and handwashing reduce the secondary attack rate by 40%, if decision-makers seek to keep the monthly probability of an in-school transmission below 50%, additional mitigation (e.g., masking) would need to be added at a community incidence of approximately 2/100,000/day. Once students are vaccinated, thresholds shift substantially higher.

**Limitations:** The interpretation of model results should be limited by the uncertainty in many of the parameters, including the effectiveness of individual mitigation interventions and vaccine efficacy against the delta variant, and the limited scope of the model beyond the school community. Additionally, the assumed case detection rate (33% of cases detected) may be too high in areas with decreased testing capacity.

**Conclusion:** Despite the assumption of high adult vaccination, the risks of both in-school SARS-CoV-2 transmission and resulting infections among students, educators/staff, and their household members remain high when the delta variant predominates and students are unvaccinated. Mitigation measures or vaccinations for students can substantially reduce these risks. These findings underscore the potential role for responsive plans, where mitigation is deployed based on local COVID-19 incidence and vaccine uptake.

## INTRODUCTION

CDC recommends in-person education for all K-12 students, with COVID-19 mitigation measures including distancing, ventilation, and indoor masking regardless of vaccination status.^1^ Vaccination is now authorized for children <12. In communities with high vaccination rates and low COVID-19 incidence, or where masking is less widely accepted, schools may remove masks and other mitigation requirements, with uncertain impact on COVID-19 outcomes for elementary students, educators/staff, and their households.

## METHODS

We used an agent-based dynamic transmission model of SARS-CoV-2 in schools. Model structure and data inputs are described in previous publications; the Supplement describes parameterization specific to this analysis.^2^ Current and prior^2^ reporting adhere to CHEERS guidelines; this was designated not human subjects research.

We simulated an elementary school (30 separate classes, 638 students, 60 educators/staff) across 12 different combinations of: three viral infectiousness levels (reflecting wild-type virus, alpha variant, and delta variant) and four student vaccination levels (0%, 25%, 50%, and 70% coverage). We assumed that 70% of adults (educators/staff and adult household members of students and educators/staff) were vaccinated (sensitivity analyses for 50% adult vaccination coverage can be found in the Supplement). For each scenario, we varied observed community COVID-19 incidence (0-50 cases/100,000 people/day, 33% of cases detected) and mitigation effectiveness (0-100% reduction to in-school secondary attack rate). Without clinical data for individual mitigation measure effectiveness, we created examples based on observational data, particle and aerosol studies, and expert opinion, to reflect three levels of in-school mitigation intensiveness: A) ventilation and handwashing only (20-40% effectiveness); B) masking plus ventilation/handwashing (70%-80%); and C) combined masking, distancing, cohorting, handwashing, and ventilation (90-100%). These ranges are highly uncertain (Supplement).

We evaluated two primary outcomes over a 30-day period: 1) probability of any in-school SARS-CoV-2 transmission with varying mitigation effectiveness, and 2) average increase in total infections among students, educators/staff, and their household members (“immediate school community”) associated with moving from more to less intensive mitigation measures (e.g., unmasking). We projected the anticipated increased in cases associated with three discrete changes in mitigation effectiveness, reflecting the midpoints or bounds of the A and B mitigation scenarios above: 70% to 40% mitigation effectiveness (smaller decrease); 75% to 35% effectiveness (moderate decrease); and 80% to 20% effectiveness (larger decrease).

## RESULTS

Over 30 days in the simulated elementary school, the probability of at least one in-school SARS-CoV-2 transmission varied widely (Figure 1). With the delta variant and no student vaccination, if decision-makers seek to keep the monthly probability of an in-school transmission below 50%, additional mitigation (e.g., masking) would need to be added to ventilation/handwashing at a community incidence of approximately 2/100,000/day, assuming 40% effectiveness of ventilation/handwashing (A) (Figure 1, top right panel, left arrow). As an alternative decision threshold, if decision-makers are willing to accept, in association with unmasking, an average of 5 additional infections per month in the immediate school community, masks could be removed at a community incidence of approximately 5/100,000/day, assuming 40% effectiveness of (A) and 70% effectiveness of (B) (Figure 2, top right panel, red line).

**Figure 1:**
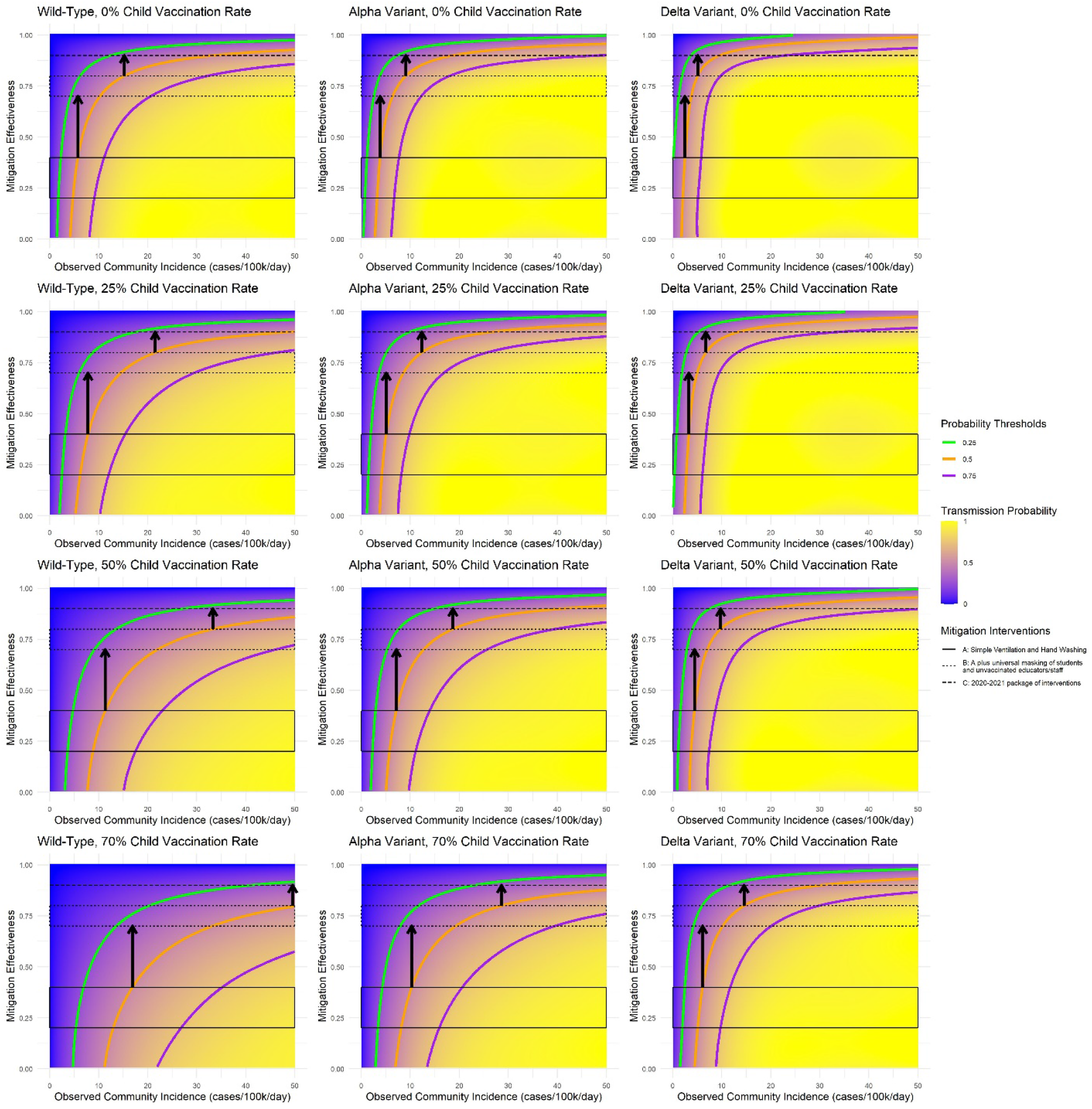
Model-estimated probability of at least one in-school SARS-CoV-2 transmission over 30 days in a simulated elementary school setting. The blue-to-yellow color scale depicts the smoothed model-estimated probability of <u>at least one in-school SARS-CoV-2 transmission over a 30-day period</u>. Panels reflect increasingly transmissible variants from left to right, and increasing student vaccination coverage from top to bottom. The horizontal axis shows observed community COVID-19 incidence in cases/100,000 people per day. The vertical axis shows mitigation effectiveness, applied as a relative risk reduction to the fully unmitigated attack rate for each variant. Bands of mitigation effectiveness reflect approximate assumptions for three types of interventions – see the Supplemental Table for more information about these effectiveness estimates. The contour lines represent thresholds for different probability levels; probabilities are lower than the threshold above the contour line and higher below it. The arrows indicate the community COVID-19 incidence rate at which a school might opt to move to the next more intensive mitigation strategy (i.e., 40% to 70% and 80% to 90% effectiveness), if the goal is to maintain the probability of the one in-school transmission per month below 50%. Adult vaccination coverage is assumed to be 70% in all scenarios (see Supplemental Figure 1 for a sensitivity analysis with 50% adult vaccination coverage).

**Figure 2:**
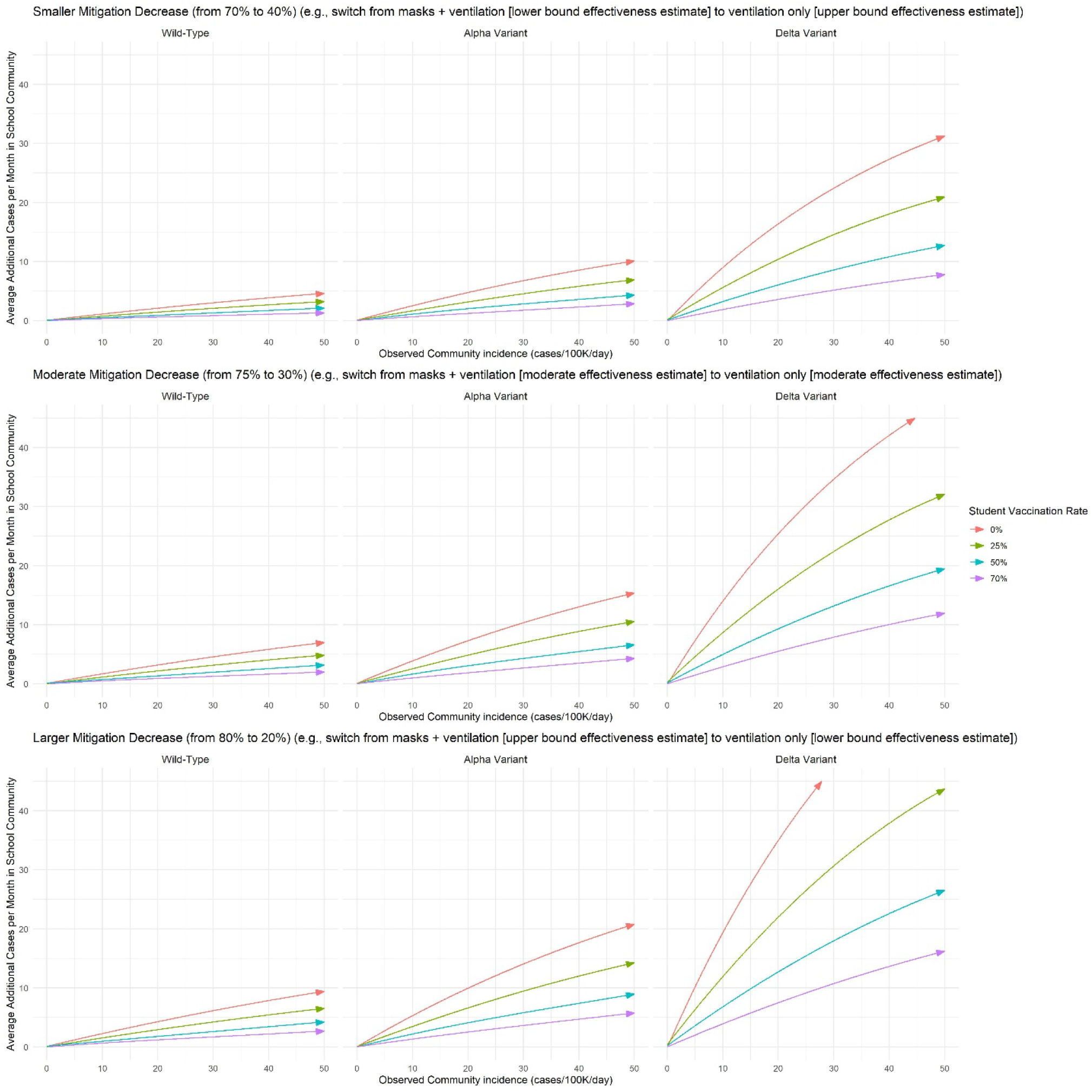
Model-estimated average number of additional cases over 30 days in the immediate school community (students, educators/staff, and their household members) associated with moving from intensive to less-intensive mitigation measures in the simulated elementary school setting. The vertical axis shows the smoothed <u>average additional number of model-estimated infections over 30 days among members of the immediate school community</u> (defined as students, educators/staff, and their household members) when moving from intensive to less intensive mitigation measures. The horizontal axis shows observed community COVID-19 incidence in cases/100,000 people per day. Panels reflect increasingly transmissible variants from left to right, and smaller differences in effectiveness between intensive and less intensive mitigation measures from top to bottom. The changes in mitigation effectiveness reflect the midpoints or bounds of the A and B mitigation scenarios presented in Figure 1: 70% to 40% mitigation effectiveness (smaller effectiveness decrease); 75% to 35% effectiveness (moderate effectiveness decrease); and 80% to 20% effectiveness (larger effectiveness decrease). Adult vaccination coverage is assumed to be 70% in all scenarios (see Supplemental Figure 2 for a sensitivity analysis with 50% adult vaccination coverage).

Increasing student vaccination to 50% in these two scenarios increased the associated thresholds to about 4 and 16 cases/100,000/day, respectively. Decreases in the assumed effectiveness of ventilation/handwashing, increases in the assumed effectiveness of masking, or decreases in the assumed proportion of all community infections that are detected shifted the thresholds lower.

## DISCUSSION

Despite high adult vaccination, the risks of in-school SARS-CoV-2 transmission and resulting infections among students, educators/staff, and their household members remain high when the delta variant predominates and students are unvaccinated. Mitigation measures or vaccinations for students can substantially reduce these risks, especially when implemented together.

Risks related to SARS-CoV-2 infection are only one of many factors guiding K-12 school planning, alongside educational, health, and social/emotional considerations. These results should be interpreted in the context of model limitations, including uncertainty in available data (e.g., effectiveness of individual mitigation interventions, case detection rate) and the focus on only the immediate school community; we excluded testing as a mitigation intervention (evaluated in previous work^3^). These findings underscore the potential role for responsive plans, where mitigation is deployed based on local COVID-19 incidence and vaccine uptake.

## Data Availability

Model code and replication files are publicly available on GitHub.

https://github.com/abilinski/BackToSchool2

## ETHICAL REVIEW

This work was designated “not human subjects research” by the Mass General Brigham Institutional Review Board (protocol 2021P002876).

## ACKNOWLEDGEMENTS

We are grateful to Dr. Sandra B. Nelson, MD (Massachusetts General Hospital) and Dr. Shira Doron, MD (Tufts Medical Center) for expert opinion on mitigation measure effectiveness. (Neither individual was compensated for their support on this work.) John Giardina had full access to all the data in the study and takes responsibility for the integrity of the data and the accuracy of the data analysis.

## SUPPLEMENT

**Supplemental Table :**
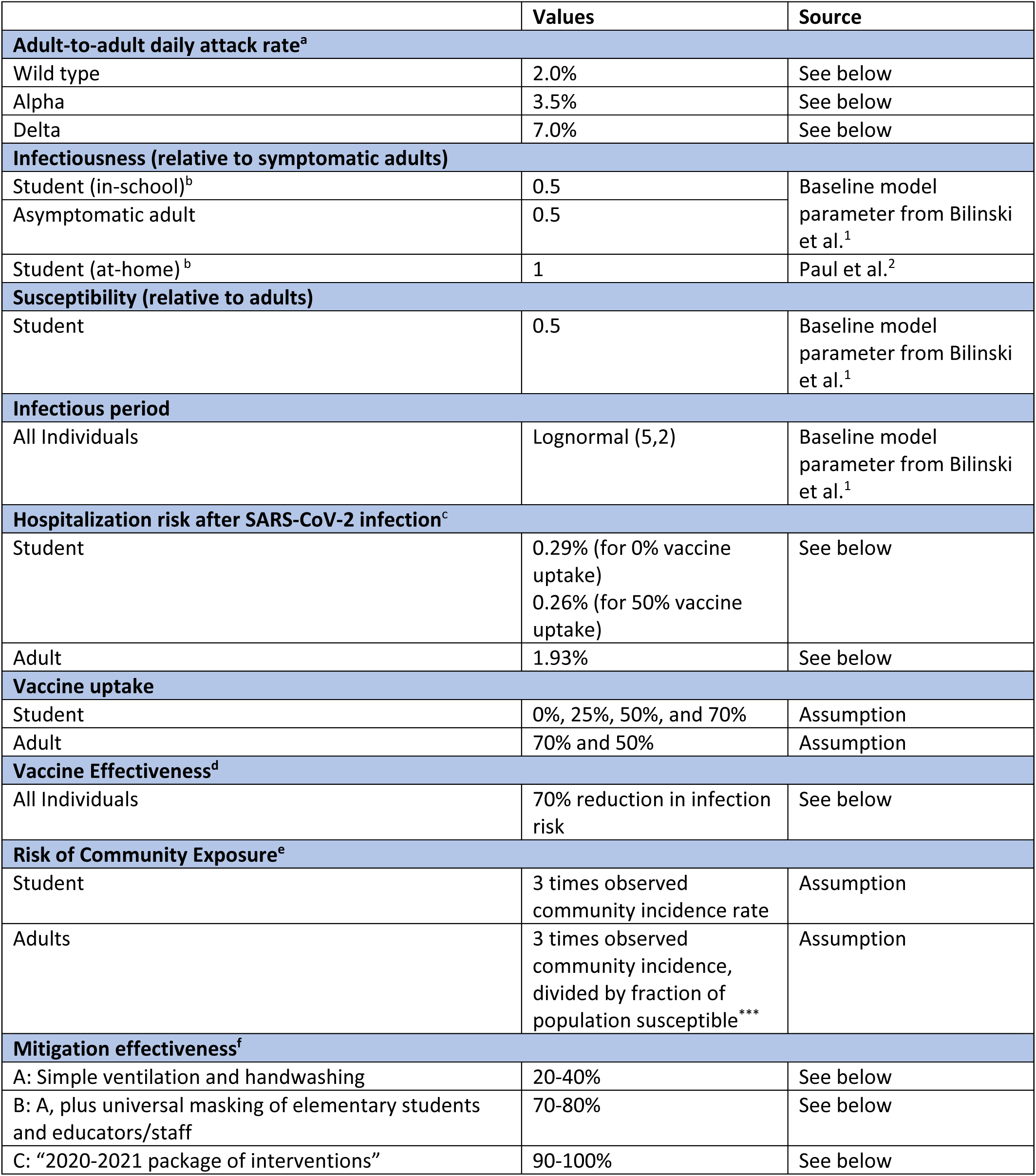
Selected input parameters for agent-based dynamic transmission model of 30-day SARS-CoV-2 outcomes in elementary schools. a. Adult-to-adult daily attack rate: Based on calibration analyses described in previous work (Bilinski et al.^1^), the adult-to-adult wild-type variant attack rate was assumed to be 2% per day. This is also consistent with data from school settings with minimal mitigation; a lower-bound estimate from CDC data suggest a total in-school attack rate of 11%,^3^ corresponding to a daily attack rate near 2% (assuming a constant daily attack rate over a 5 day infectiousness period). The transmissibility of the alpha variant is estimated at 59% higher than wild type in the US,^4^ so we assumed a daily attack rate of 3.5%. The delta attack rate has been estimated at 60% higher than that for the alpha variant in a setting with high vaccine uptake,^5^ and a case study of a recent outbreak in a largely unvaccinated population reported an overall household delta attack rate of 53%,^6^ corresponding to a daily attack rate of approximately 7% per school day (again assuming a 5 day infectiousness period and an in-school attack rate approximately half the household attack rate). In order to reflect the baseline attack rate, the estimate from the largely unvaccinated population was used. The student-to-student daily attack rate was assumed to be one-quarter of the adult-to-adult rate to reflect the assumption that students are both 50% less infectious and 50% less susceptible to infection. b. Infectiousness of students: Based on a review of the evidence by Bilinski et al.^1^, it was assumed that elementary school students are half as infectious as adults in a school setting. A recent study from Paul et al.^2^, however, suggests the transmission dynamics within a household may be different from what has been previously observed in a school setting – their findings suggest that younger children are more likely to infect others in a household than older children, potentially due to how parents interact with children of that age. To incorporate this additional evidence, elementary school aged children were assumed to be as infectious as adults within the household setting. c. Hospitalization risk following SARS-CoV-2 infection: The hospitalization risk among unvaccinated patients with SARS-CoV-2 was assumed to be the overall infection fatality rate divided by the mortality rate among hospitalized patients, using estimates provided by the CDC for use in COVID-19 models^7^; the 0-17 year old age group (20/million IFR; 0.7% hospitalized mortality rate) was used for the student estimate and the 18-49 year old age group (500/million IFR; 2.1% hospitalized mortality rate) was used for the adult estimate. The overall hospitalization risk among both unvaccinated and vaccinated patients was calculated using the vaccine coverage scenarios implemented in the model and assuming that the risk of infection in vaccinated individuals was 90% lower than the risk in unvaccinated individuals. Adult values applied to educators/staff and to adult household members of both students and educators/staff. The 2% hospitalization rate used for adults is similar to inputs in other models, such as the low estimate used by Lemaitre et al.^8^ d. Vaccine effectiveness: There is significant uncertainty around vaccine effectiveness after accounting for the potential for waning immunity and the relatively recent predominance of the delta variant. Since results from the initial clinical trials for the vaccines do not account for waning efficacy (beyond their initial study period) and the emergence of the delta variant, more recent observational data were used to parameterize the model. Some recent estimates of vaccine effectiveness against infection from observational studies in the US covering the months where delta was dominant include: 75% among New York state residents identified through vaccine and testing registries;^9^ 66% among healthcare workers in a California health system;^10^ 66% among frontline workers participating in a prospective cohort study;^11^ and 76% and 42% for the mRNA-1273 and BNT162b2 vaccines, respectively, among patients receiving testing through the Mayo Clinic Health System.^12^ A broader pre-print meta-analysis on vaccine effectiveness also estimated an effectiveness of 72% delta.^13^ These findings led to our assumption of 70% vaccine effectiveness against infection. It should be noted, however, that it is not necessarily clear how generalizable the existing empirical data is to elementary school-age children, especially since children have previously been found to be less susceptible to the virus and will be newly vaccinated following approval for their age group (and will not have had time to experience waning immunity). Since there is no reliable evidence on the effect of these factors, however, the assumption was maintained at 70%. If effectiveness in children is higher than 70%, there will generally be lower transmission within schools than predicted by the model, and vice versa. (The higher vaccine coverage scenarios may provide insight into the extent of the impact of vaccine efficacy -- for example, in the context of the model, the 70% coverage scenario with 70% effectiveness would be roughly equivalent to a scenario with 50% coverage but almost 100% effectiveness.) e. Risk of community exposure was inflated by the fraction of population susceptible (i.e., unvaccinated and potential breakthrough cases), so that actual case rate among adults matches the assumed community incidence rate. f. Mitigation measures: **A.** Simple ventilation and handwashing: open windows if present, portable air filters, maintain existing HVAC systems, and regular handwashing. Vouriot et al.^14^ estimate that seasonal changes in ventilation increase secondary infections by 30-40% in fall and 80-90% in winter relative to summer, but note there is wide variation based on classroom activities. If an intervention replicates summer-levels of ventilation, this would correspond to a 23-29% reduction in the attack rate in the fall and a 44-47% reduction in the winter. Burridge et al.^15^ also present a wide range of studies on ventilation and surface cleaning, some of which show good ventilation (i.e., opening windows) could reduce the risk of infections by about half in an office setting (which is often less active than a classroom). Data from airflow studies estimate reduction in exposure to aerosols of 65% with portable HEPA filters.^16^ Combining these data, we estimated a range of 20-40% risk reduction (since most classrooms will not have access to portable air cleaners) **B.** Interventions in A, plus universal masking: a policy of masking all students and educators/staff, accounting for the impact of imperfect adherence and technique. There are few studies on the specific combination of masking and ventilation/handwashing that do not consider other interventions as well. Considering data from studies of masks alone, a meta-analysis estimated that using non-medical masks was, depending on the model used, associated with a 43% or 35% reduction in respiratory virus infection risk.^17^ A recent working paper from Abaluck et al.^18^ found that a cluster randomized intervention in Bangladesh to promote the use of masks increased mask use by 29% and was associated with a 9.3% percentage point decrease in COVID-19 seroprevalence. The authors note that the effect of universal masking would likely be several times higher; a rough, back-of-the-envelope rescaling of this effect using a basic Wald estimator to evaluate the impact of universal masking provides an estimate of a 32% (9.3%/29%) reduction in seroprevalence from universal masking, which is in line with the meta-analysis estimates on the reduction in infection risk from masking alone. Similarly, a meta-analysis on mask effectiveness across studies focusing on SARS-CoV-1, MERS, and SARS-CoV-2 by Chu et al.^19^ found a 44% reduction in risk for masks used in the community. There is substantial uncertainty in this estimate, with many studies reporting higher risk reductions.^20^ Additionally, data from studies of simulated respiratory particles demonstrate fitted filtration efficiency values (proportion of particles kept behind a mask) of 50-79% with cloth masks.^21,22^ An experimental and mathematical modeling study on aerosol dynamics from Rothamer et al.^23^ found that a non-medical procedure mask with a baseline effectiveness of 29% had an effectiveness of 62- 77% when combined with different levels of ventilation, with this combined effectiveness increasing for masks with better baseline effectiveness. This is also consistent with the meta-analysis from Chu et al.^19^ on masking in a healthcare setting (likely to be accompanied by ventilation; 70% reduction in risk) and a range of case studies suggesting masking effectiveness can reach as high as 70-79% in non-school settings, including in some households in China and during an outbreak on a US Navy ship.^20^ We anticipated that the combined effectiveness of both masking and ventilation/handwashing (B) would be between the effectiveness estimates for A and C, and based on the data above, we assumed that masking and ventilation/handwashing was approximately 70-80% effective. **C.** Combination interventions as implemented in many settings in the 2020-2021 school year: Includes B, plus physical distancing of 3-6 feet when masked and >6 when unmasked, daily cleaning of surfaces, restrictions on shared items, and cohorting of students.^24^ This is the assumed maximum mitigation effect. CDC reports very effective in-school mitigation when the full package of interventions are implemented,^24^ including those from Falk et al.^25^ and Zimmerman et al.^26^ Many studies reported total in-school secondary attack rates of 0.5-1.0% with implementation of this package of interventions; this corresponds to a 93% reduction on the unmitigated wild-type SAR. While clinical data are available for C, data about the effectiveness of A and B were derived from the limited data cited above, as well as expert opinion, and should be used only as approximations to estimate the impact of individual mitigation interventions. Model code is available at: https://github.com/abilinski/BackToSchool2/tree/master/3 - Scripts/Paper 3

|  | Values | Source |
| --- | --- | --- |
| **Adult-to-adult daily attack rate<sup>a</sup>** |  |  |
| Wild type | 2.0% | See below |
| Alpha | 3.5% | See below |
| Delta | 7.0% | See below |
| **Infectiousness (relative to symptomatic adults)** |  |  |
| Student (in-school)<sup>b</sup> | 0.5 | Baseline model parameter from Bilinski et al.<sup>1</sup> |
| Asymptomatic adult | 0.5 |  |
| Student (at-home)<sup>b</sup> | 1 | Paul et al.<sup>2</sup> |
| **Susceptibility (relative to adults)** |  |  |
| Student | 0.5 | Baseline model parameter from Bilinski et al.<sup>1</sup> |
| **Infectious period** |  |  |
| All Individuals | Lognormal (5,2) | Baseline model parameter from Bilinski et al.<sup>1</sup> |
| **Hospitalization risk after SARS-CoV-2 infection<sup>c</sup>** |  |  |
| Student | 0.29% (for 0% vaccine uptake)<br>0.26% (for 50% vaccine uptake) | See below |
| Adult | 1.93% | See below |
| **Vaccine uptake** |  |  |
| Student | 0%, 25%, 50%, and 70% | Assumption |
| Adult | 70% and 50% | Assumption |
| **Vaccine Effectiveness<sup>d</sup>** |  |  |
| All Individuals | 70% reduction in infection risk | See below |
| **Risk of Community Exposure<sup>e</sup>** |  |  |
| Student | 3 times observed community incidence rate | Assumption |
| Adults | 3 times observed community incidence, divided by fraction of population susceptible *** | Assumption |
| **Mitigation effectiveness<sup>f</sup>** |  |  |
| A: Simple ventilation and handwashing | 20-40% | See below |
| B: A, plus universal masking of elementary students and educators/staff | 70-80% | See below |
| C: "2020-2021 package of interventions" | 90-100% | See below |

### Smoothing Methods for Figures

To generate Figures 1 and 2 and Supplemental Figures 1 and 2, for each scenario we ran the 100 replicates of the model presented in Bilinski et al.^1^ for each combination of observed community incidence from 0 to 60 case notifications/100k/day (incremented by 1) and mitigation effectiveness from 0 to 100% (incremented by 1%) using the model code available at https://github.com/abilinski/BackToSchool2/tree/master/3-Scripts/Paper3. The scenarios included combinations of the following parameter values: (1) baseline daily adult-to-adult secondary attack rates of 2% (reflecting the wild-type virus), 3.5% (alpha variant), and 7% (delta variant); (2) child vaccination coverage of 0%, 25%, 50%, and 70%; and (3) adult vaccination coverage of 50% and 70%. (The 70% child vaccination scenario was only run when adult vaccination coverage was 70% as well, to reflect observed patterns of vaccine uptake.) We assumed reduced transmission and susceptibility for children and for asymptomatic adults (Supplemental Table).

The raw model output is highly stochastic, so to generate the smoothed heatmaps and associated contour line estimates in Figure 1 and the smoothed line graphs in Figure 2, we fit regressions for each outcome and associated scenario (e.g., more than one in-school transmission in the wild-type, 0% child vaccine, 70% adult vaccine scenario) as a function of observed community incidence and mitigation. The outcome value used to fit each regression was the mean of 90 replicates in a training sample at each combination of incidence and mitigation. (Note that for the additional cases metric in Figure 2, we fit the regression to the overall number of cases in the immediate school community, and then subtracted the fitted regression across the different mitigation levels to generate the estimated average additional cases from moving between each mitigation level.) We tested five specifications: linear, quadratic, cubic, and quartic polynomials, as well as linear regression with a log transformation on each predictor:

- Linear specification: *outcome*=*β*_0_ + *β*_1_*Incidence* + *β*_2_ *Mitigation*+ *β*_3_*Incidence *Mitigation*
- Quadratic specification: 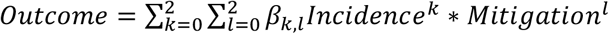
- Cubic specification: 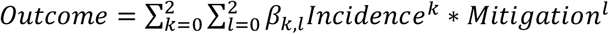
- Quartic specification: 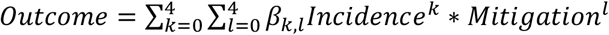
- Log specification: *outcome*=*β*_0_ + *β*_1_ In *Incidence*+ *β*_2_ In *Mitigation*+ *β*_3_ In *Incidence\** In *Mitigation*

For each scenario, we selected the regression which minimized the root mean-squared prediction error in a hold-out test set containing 10 replicates (10%) at each combination of incidence and mitigation.

To assess how well the smoothing functions fit the expected value of the model output, we calculated the R^2^ between binned averages of the model-generated outcomes in the hold-out test set and the average outcome predicted by the selected smoothing function across the range of community incidence and mitigation values. We evaluated the fit for two different bin sizes: “large” bins, with a bin width of 5 for community incidence and 0.1 for mitigation effectiveness, and “small” bins with a bin width of 1 for community incidence and 0.1 for mitigation effectiveness. The lowest R^2^ was 0.94 for the small bins and 0.98 for the large bins, indicating that the smoothing procedure to generate the figures accurately reflects the average model output over the different scenarios analyzed within these bin sizes.

**Supplemental Figure 1:**
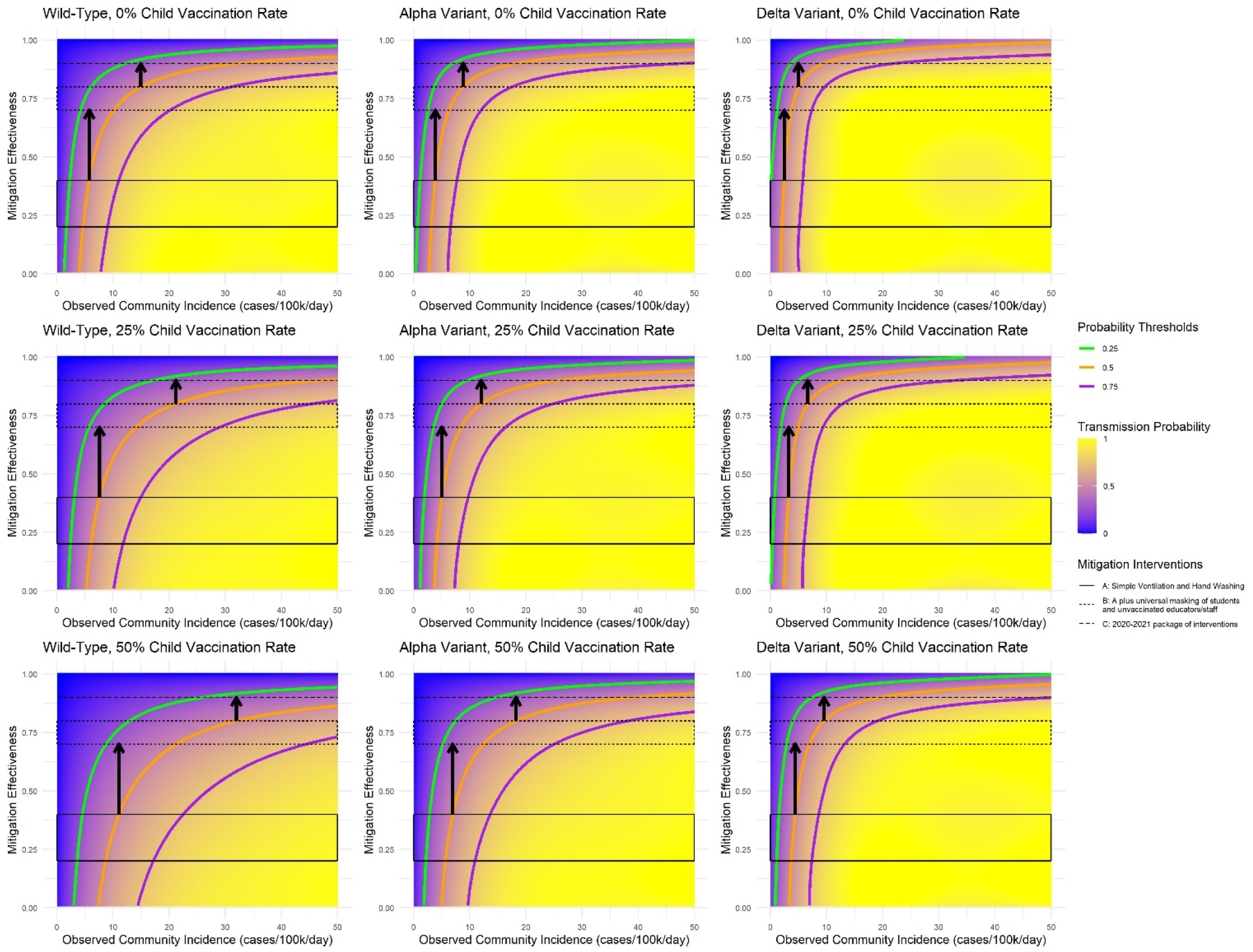
Model-predicted probability of at least one in-school SARS-CoV-2 transmission over 30 days in a simulated elementary school setting, 50% adult vaccination coverage.

**Supplemental Figure 2:**
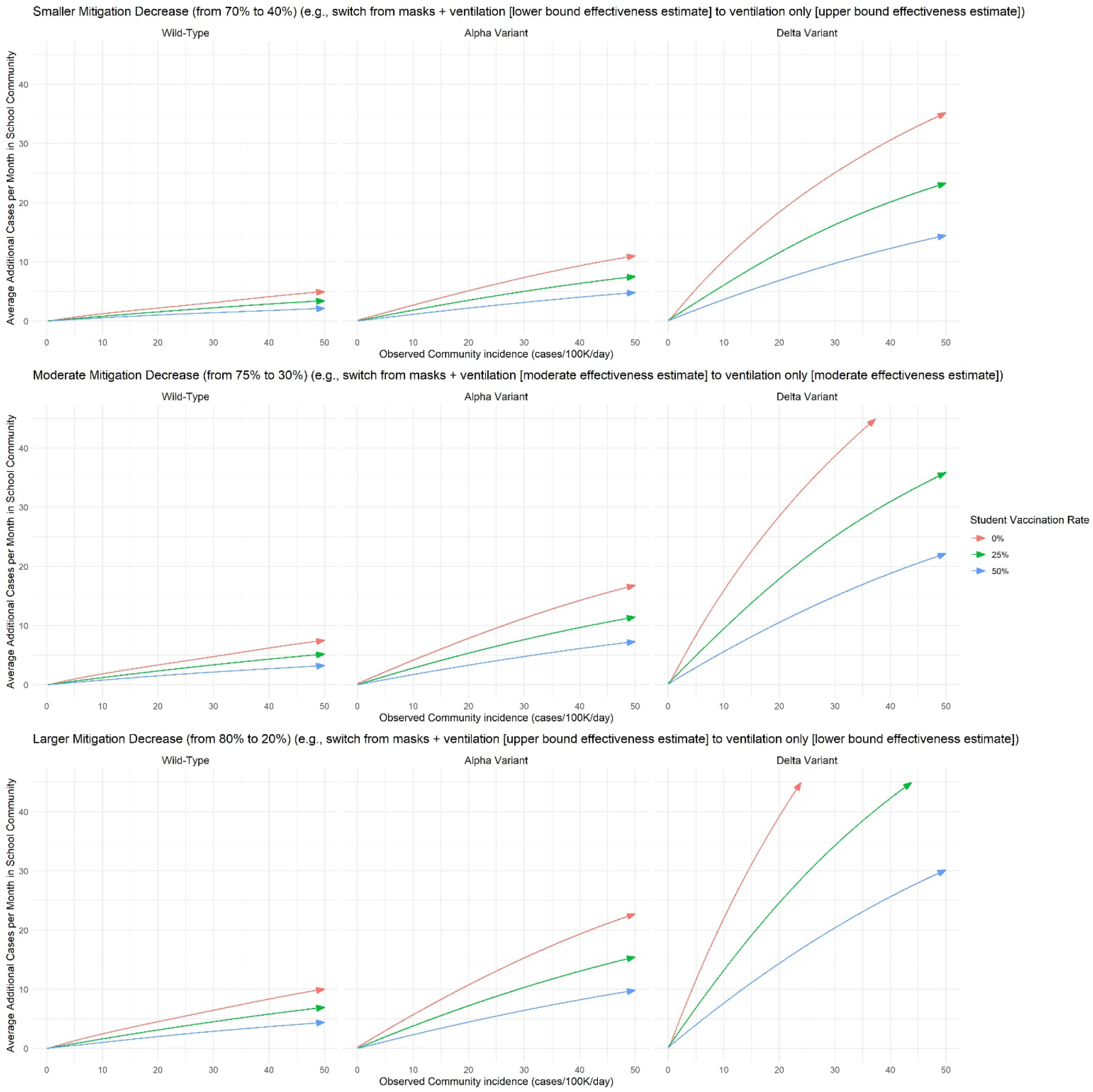
Model-estimated average number of additional cases over 30 days in the immediate school community (students, educators/staff, and their household members) associated with moving from intensive to less-intensive mitigation measures in the simulated elementary school setting, 50% adult vaccination coverage.

**Supplemental Figure 3:**
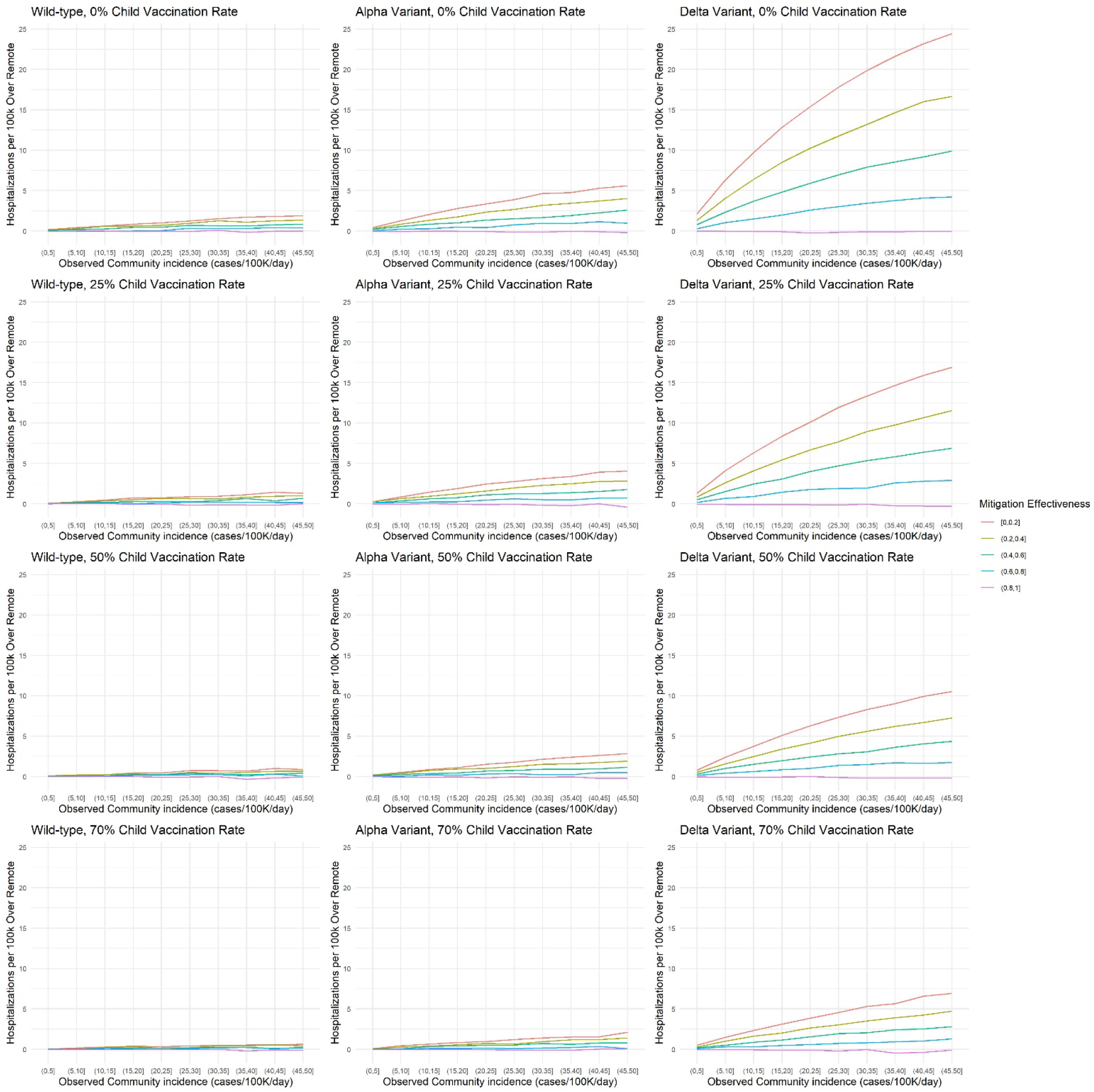
Average increase in hospitalizations among parents, teachers, staff, and adult family members per 100k, relative to remote instruction, with 70% adult vaccination.

**Supplemental Figure 4:**
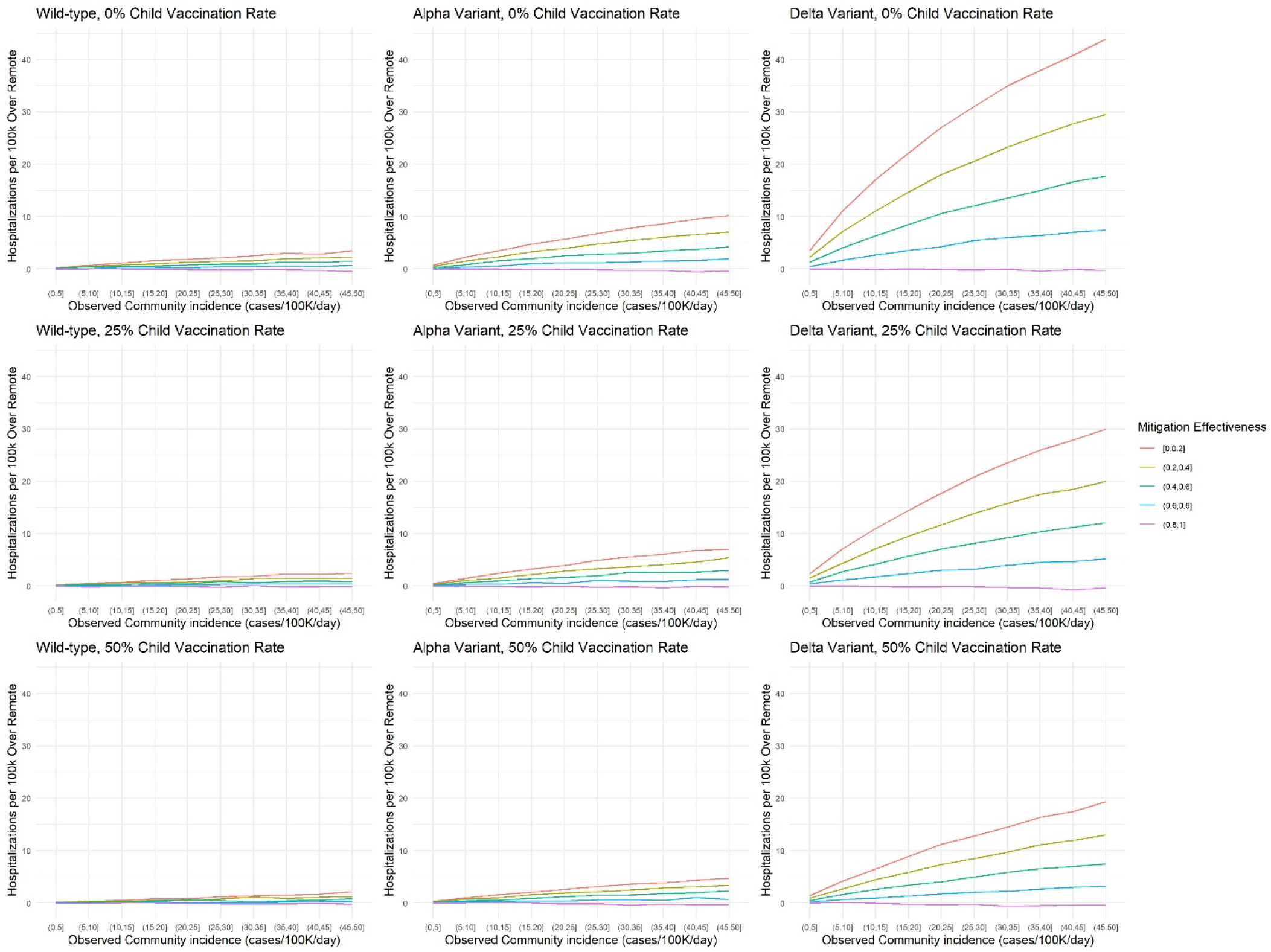
Average increase in hospitalizations among parents, teachers, staff, and adult family members per 100k, relative to remote instruction, with 50% adult vaccination.

## REFERENCES

1. Centers for Disease Control and Prevention. Guidance for COVID-19 Prevention in K-12 Schools. Published July 9, 2021. Accessed July 16, 2021. https://www.cdc.gov/coronavirus/2019-ncov/community/schools-childcare/k-12-guidance.html

2. Bilinski A, Salomon JA, Giardina J, Ciaranello A, Fitzpatrick MC. Passing the Test: A Model-Based Analysis of Safe School-Reopening Strategies. Ann Intern Med. Published online June 8, 2021:M21-0600. doi:10.7326/M21-0600

3. Bilinski A, Ciaranello A, Fitzpatrick MC, et al. SARS-CoV-2 testing strategies to contain school-associated transmission: model-based analysis of impact and cost of diagnostic testing, screening, and surveillance. medRxiv. 2021.05.12.21257131. doi:10.1101/2021.05.12.21257131

## REFERENCES

1. Bilinski A, Salomon JA, Giardina J, Ciaranello A, Fitzpatrick MC. Passing the Test: A Model-Based Analysis of Safe School-Reopening Strategies. Ann Intern Med. Published online June 8, 2021:M21-0600. doi:10.7326/M21-0600

2. Paul LA, Daneman N, Schwartz KL, et al. Association of Age and Pediatric Household Transmission of SARS-CoV-2 Infection. JAMA Pediatr. Published online August 16, 2021. doi:10.1001/jamapediatrics.2021.2770

3. Doyle T, Kendrick K, Troelstrup T, et al. COVID-19 in Primary and Secondary School Settings During the First Semester of School Reopening — Florida, August–December 2020. MMWR Morb Mortal Wkly Rep. 2021;70(12):437–441. doi:10.15585/mmwr.mm7012e2

4. Davies NG, Abbott S, Barnard RC, et al. Estimated transmissibility and impact of SARS-CoV-2 lineage B.1.1.7 in England. Science. 2021;372(6538):eabg3055. doi:10.1126/science.abg3055

5. Public Health England. SARS-CoV-2 Variants of Concern and Variants under Investigation in England.; 2021. https://assets.publishing.service.gov.uk/government/uploads/system/uploads/attachment_data/file/990339/Variants_of_Concern_VOC_Technical_Briefing_13_England.pdf

6. Dougherty K, Mannell M, Naqvi O, Matson D, Stone J. SARS-CoV-2 B.1.617.2 (Delta) Variant COVID-19 Outbreak Associated with a Gymnastics Facility — Oklahoma, April–May 2021. MMWR Morb Mortal Wkly Rep. 2021;70(28):1004–1007. doi:10.15585/mmwr.mm7028e2

7. Centers for Disease Control and Prevention. COVID-19 Pandemic Planning Scenarios. Published March 19, 2021. Accessed July 27, 2021. https://www.cdc.gov/coronavirus/2019-ncov/hcp/planning-scenarios.html

8. Lemaitre JC, Grantz KH, Kaminsky J, et al. A scenario modeling pipeline for COVID-19 emergency planning. Sci Rep. 2021;11(1):7534. doi:10.1038/s41598-021-86811-0

9. Rosenberg ES, Holtgrave DR, Dorabawila V, et al. New COVID-19 Cases and Hospitalizations Among Adults, by Vaccination Status — New York, May 3–July 25, 2021. MMWR Morb Mortal Wkly Rep. 2021;70(37):1306–1311. doi:10.15585/mmwr.mm7037a7

10. Keehner J, Horton LE, Binkin NJ, et al. Resurgence of SARS-CoV-2 Infection in a Highly Vaccinated Health System Workforce. N Engl J Med. 2021;385:1330–1332. doi:10.1056/NEJMc2112981

11. Fowlkes A, Gaglani M, Groover K, et al. Effectiveness of COVID-19 Vaccines in Preventing SARS-CoV-2 Infection Among Frontline Workers Before and During B.1.617.2 (Delta) Variant Predominance — Eight U.S. Locations, December 2020–August 2021. MMWR Morb Mortal Wkly Rep. 2021;70(34):1167–1169. doi:10.15585/mmwr.mm7034e4

12. Puranik A, Lenehan PJ, Silvert E, et al. Comparison of two highly-effective mRNA vaccines for COVID-19 during periods of Alpha and Delta variant prevalence. medRxiv. 2021.08.06.21261707. doi:10.1101/2021.08.06.21261707

13. Zeng B, Gao L, Zhou Q, et al. Effectiveness of COVID-19 vaccines against SARS-CoV-2 variants of concern: a systematic review and meta-analysis. medRxiv. 2021.09.23.21264048. doi:10.1101/2021.09.23.21264048

14. Vouriot CVM, Burridge HC, Noakes CJ, Linden PF. Seasonal variation in airborne infection risk in schools due to changes in ventilation inferred from monitored carbon dioxide. Indoor Air. 2021;31(4):1154–1163. doi:10.1111/ina.12818

15. Burridge HC, Bhagat RK, Stettler MEJ, et al. The ventilation of buildings and other mitigating measures for COVID-19: a focus on wintertime. Proc R Soc Math Phys Eng Sci. 2021;477(2247):20200855. doi:10.1098/rspa.2020.0855

16. Lindsley WG, Derk RC, Coyle JP, et al. Efficacy of Portable Air Cleaners and Masking for Reducing Indoor Exposure to Simulated Exhaled SARS-CoV-2 Aerosols — United States, 2021. MMWR Morb Mortal Wkly Rep. 2021;70(27):972–976. doi:10.15585/mmwr.mm7027e1

17. Reiner RC, Barber RM, Collins JK, et al. Modeling COVID-19 scenarios for the United States. Nat Med. 2021;27(1):94–105. doi:10.1038/s41591-020-1132-9

18. Abaluck J, Kwong LH, Styczynski A, et al. The Impact of Community Masking on COVID-19: A Cluster-Randomized Trial in Bangladesh. Innovations for Poverty Action. 2021. Accessed October 11, 2021. https://www.poverty-action.org/publication/impact-community-masking-covid-19-cluster-randomized-trial-bangladesh

19. Chu DK, Akl EA, Duda S, et al. Physical distancing, face masks, and eye protection to prevent person-to-person transmission of SARS-CoV-2 and COVID-19: a systematic review and meta-analysis. Lancet. 2020;395(10242):P1973–1987. doi:10.1016/S0140-6736(20)31142-9

20. Brooks JT, Butler JC. Effectiveness of Mask Wearing to Control Community Spread of SARS-CoV-2. JAMA. 2021;325(10):998. doi:10.1001/jama.2021.1505

21. Centers for Disease Control and Prevention. Science Brief: Transmission of SARS-CoV-2 in K-12 Schools and Early Care and Education Programs. Centers for Disease Control and Prevention. Published July 9, 2021. Accessed July 28, 2021. https://www.cdc.gov/coronavirus/2019-ncov/science/science-briefs/transmission_k_12_schools.html

22. Clapp PW, Sickbert-Bennett EE, Samet JM, et al. Evaluation of Cloth Masks and Modified Procedure Masks as Personal Protective Equipment for the Public During the COVID-19 Pandemic. JAMA Intern Med. 2021;181(4):463. doi:10.1001/jamainternmed.2020.8168

23. Rothamer DA, Sanders S, Reindl D, Bertram TH. Strategies to minimize SARS-CoV-2 transmission in classroom settings: combined impacts of ventilation and mask effective filtration efficiency. Science and Technology for the Built Environment. 2021;27(9):1181–1203. doi:10.1080/23744731.2021.1944665

24. Centers for Disease Control and Prevention. Guidance for COVID-19 Prevention in K-12 Schools. Published July 9, 2021. Accessed July 16, 2021. https://www.cdc.gov/coronavirus/2019-ncov/community/schools-childcare/k-12-guidance.html

25. Falk A, Benda A, Falk P, Steffen S, Wallace Z, Høeg TB. COVID-19 Cases and Transmission in 17 K–12 Schools — Wood County, Wisconsin, August 31–November 29, 2020. MMWR Morb Mortal Wkly Rep. 2021;70(4):136–140. doi:10.15585/mmwr.mm7004e3

26. Zimmerman KO, Akinboyo IC, Brookhart MA, et al. Incidence and Secondary Transmission of SARS-CoV-2 Infections in Schools. Pediatrics. 2021;147(4). doi:10.1542/peds.2020-048090

